# Non-causal association between α-klotho and human lifespan: evidence from multi-omics insights

**DOI:** 10.1101/2025.02.08.25321911

**Authors:** Xueyi Liu, Wulin Yang

## Abstract

While genetic evidence robustly associates longevity in non-human primates with Klotho protein, such a direct correlation in humans remains elusive. To scrutinize the potential causal link between genetically predicted Klotho levels and human lifespan, we devised a meticulous two-sample Mendelian randomization (MR) analysis, just leveraging single nucleotide polymorphisms (SNPs) from genome-wide association studies (GWAS) and protein quantitative trait loci (pQTLs) as instrumental variables, meticulously analyzing the relationship between serum α-klotho and human longevity. By integrating MR estimates across diverse data sources using the fixed-effects inverse-variance weighted (IVW) approach, we consolidated our findings with a fixed-effects meta-analysis, fortified by sensitivity analyses embracing the simple median, weighted median, MR-Egger regression, and MR-pleiotropy residual sum and outlier assessments. Surprisingly, our genome-wide association MR analyses failed to uncover a causal association between Klotho and human lifespan, holding for both experimental and validation cohorts. Furthermore, the analysis grounded in protein quantitative trait loci also yielded no evidence of a causal link, with the sensitivity analyses consistently reinforcing the robustness of our findings. Hence, while animal models suggest a correlation between circulating Klotho and lifespan, this study demonstrates that genetically predicted levels of circulating α klotho do not exhibit a direct causal effect on human longevity.

## Introduction

In the context of global aging, the escalating prevalence of chronic diseases and soaring healthcare costs highlight the urgency to fortify preventative measures(Levine et al., 2018), While chronological age, fixed by the passage of years, is a key predictor of chronic disease risk, biological age—a dynamic reflection of physiological wear and tear—can vary widely among individuals of the same age(Wang et al., 2023). Although PhenoAge(Wang, et al., 2023) and GrimAge(Kong et al., 2023), based on biomarkers and epigenetic clocks respectively, estimate biological age, their direct correlation with lifespan prediction remains limited. Klotho, an emerging longevity biomarker, promises to revolutionize the field by offering a potential tool for predicting human lifespan.

Early research showed Klotho overexpression extended mouse lifespan, while deficiency shortened it by approximately a quarter(Xu & Sun, 2015). The *Klotho* gene encodes a kidney-predominant transmembrane protein(Donate-Correa, Martín-Carro, Cannata-Andía, Mora-Fernández, & Navarro-González, 2023), also found in the brain, parathyroid glands, peripheral blood cells, and vascular tissues(Arroyo et al., 2022). It is noteworthy that the Klotho gene family comprises three homologs: α-klotho, β-klotho, and γ-klotho, each with distinct roles(Espuch-Oliver et al., 2022). This paper focuses on α-klotho (henceforth “Klotho”) due to its direct link with aging. It inhibits aging-related signaling pathways like TGF-β, IGF-1, NF-κB, and Wnt/β-catenin(Prud’homme, Kurt, & Wang, 2022), exerting potent anti-aging effects.

Despite gene evidence suggesting a strong correlation between longevity in non-human primates and Klotho(Castner et al., 2023), a definitive genetic link between Klotho and human longevity remains elusive. Mendelian Randomization (MR), a “natural experiment“(Ference, Holmes, & Smith, 2021), effectively controls for biases in causal inference(Sekula, Del Greco, Pattaro, & Köttgen, 2016). In this study, we employ a multi-omics MR analysis to unravel the causal relationship between Klotho and human lifespan, aiming to provide a more robust and unbiased understanding of this complex interplay.

## MATERIALS AND METHODS

### Data Sources

In this study, single-nucleotide polymorphisms (SNPs) derived from Genome-wide association Analysis (GWAS) and Protein Quantitative Trait Loci (pQTL) associated with plasma Klotho levels were used as instrumental variables for MR analysis. In MR analyses based on GWAS-associated SNPs, IVs associated with exposure were derived from the studies of Ludwigshafen Risk and Cardiovascular Health (LURIC) and the Avon Longitudinal Study of Parents and Children (ALSPAC). Two studies were set, respectively, as an experimental group (n=4376) and a validation group (n=4675) (Supplementary Table 1)(Gergei et al., 2022). The pQTL data were obtained from a study conducted by a joint team from deCODE Genetics and the University of Iceland (https://www.decode.com/summarydata/). SNPs located in or close to their coding gene (cis), were selected as instrumental variables(Eldjarn et al., 2023; Sun, Chen, He, & Zheng, 2022) (Supplementary Table 2). In addition, the IVs association with the outcome (human lifespan) was obtained from four independent data sets (Supplementary Table 1).

### Instrumental SNPs Selection

The selection criteria for IVs in this study were that the SNP associated with α-klotho was at the genome-wide significance level (P<5.0×10^-8^) and was not in linkage disequilibrium (LD) with other SNPs (r^2^ < 0.3 within a clumping window of 100 kb). The F statistic was used to evaluate the strength of the IVs. A cutoff of 10 was used to distinguish between strong and weak instruments (Supplementary Table 2).

### Statistical Analysis

To ensure the robustness of the results, the inverse variance weighted (IVW) model was chosen as the main analysis method, and additional sensitivity analysis methods, MR-Egger, weighted median, and weighted mode. Heterogeneity was tested using MR Egger and IVW methods. Cochrane’s Q value was used to evaluate the heterogeneity of genetic tools. The horizontal pleiotropy of IVs was evaluated by the MR Egger regression equation. A leave-one-out analysis was performed to assess whether bias existed due to individual SNP independently affecting the results.

## RESULTS

### MR analysis based on GWAS-associated SNPs

The comprehensive MR analysis, using GWAS-linked SNPs, unveiled a consistent absence of a causal nexus between α-klotho and human lifespan in both the experimental and validation cohorts (Figure 1A & 1B). Specifically, utilizing the IVW method, the results indicated neutrality, with odds ratios (ORs) hovering around unity and non-significant P-values (OR = 1.00, 95% CI: 0.98-1.01, P = 0.46 for the experimental group, and OR = 0.99, 95% CI: 0.98-1.01, P = 0.38 for the validation set). Additional analyses within these groups further reinforced this non-causality, with ORs ranging from 0.98 to 1.02, all falling within non-significant thresholds.

**Figure 1.**
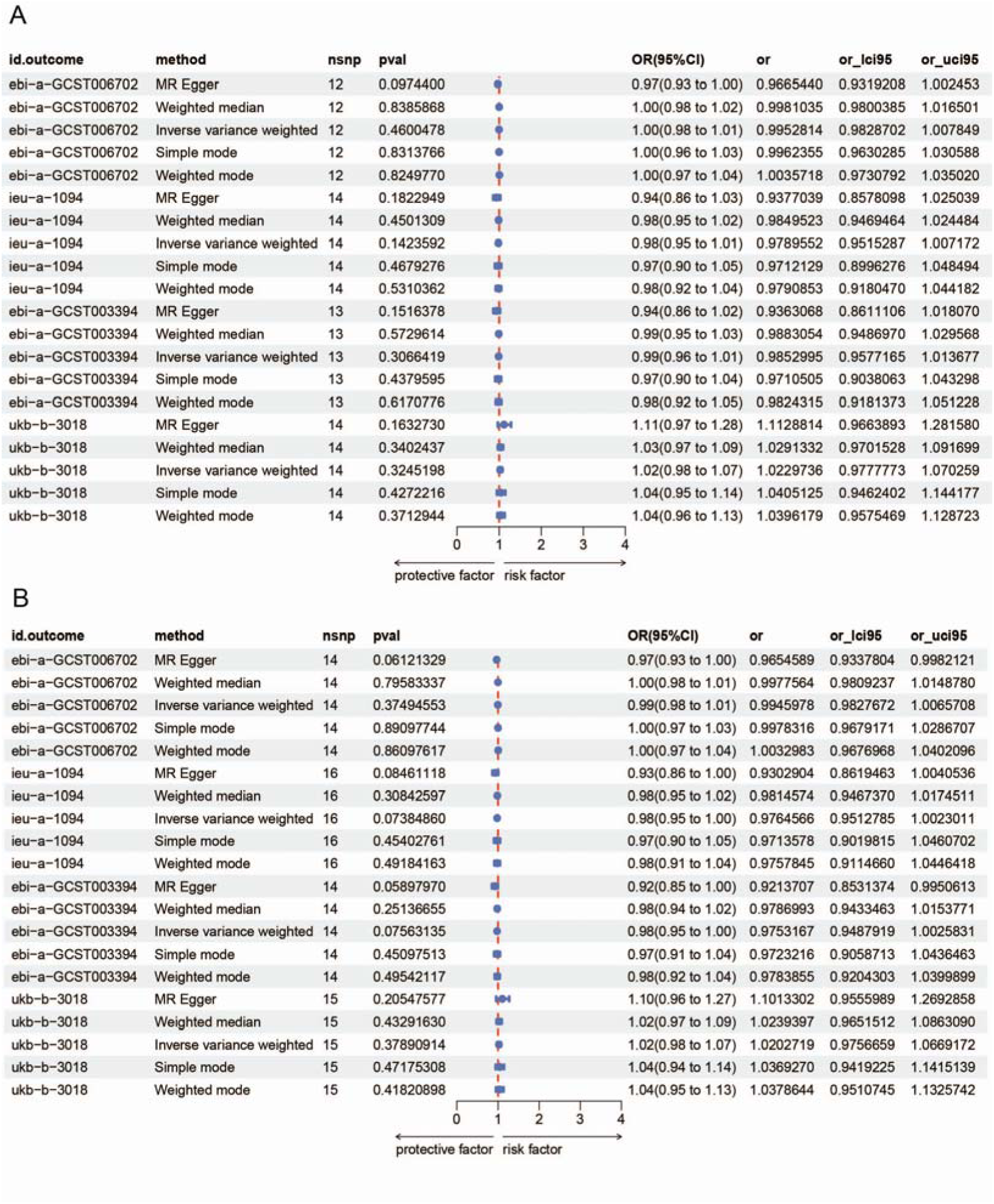
Genetic causal associations between Klotho and human lifespan based on GWAS-linked MR analysis. (A) The genetic association was estimated by MR in the experimental group. (B) The genetic association was estimated by MR in the validation cohort. The inverse variance weighted method is considered the main method.

In-depth sensitivity testing, presented in Supplementary Figure 1, failed to detect any evidence of pleiotropy in either cohort, ensuring the robustness of our findings. Furthermore, no individual SNP was identified as a solitary influencer capable of altering the overall outcome, as evident from Supplementary Figure 2. The MR-Egger Intercept analysis, designed to uncover potential horizontal pleiotropy between α-klotho and human longevity, likewise returned inconsequential results, underscoring the absence of such associations (Supplementary Table 3).

Finally, heterogeneity assessments conducted through both MR-Egger and IVW tests yielded Q values that did not attain statistical significance (all P values > 0.05), reinforcing the homogeneity and credibility of our analytical framework (Supplementary Table 4). In summary, this rigorous MR analysis, grounded in GWAS-associated SNPs, conclusively establishes the absence of a causal relationship between α-klotho and human lifespan.

### MR analysis based on pQTL-linked SNPs

Expanding on our findings, the pQTL-based MR analysis, an additional layer of scrutiny, likewise failed to uncover any causal link between α-klotho and human lifespan (Figure 2). Employing the IVW method, the results remained consistent, with odds ratios (ORs) tightly clustered around unity and accompanied by non-significant P-values (OR = 1.00, 95% CI: 0.98-1.01, P = 0.46; OR = 0.98, 95% CI: 0.95-1.02, P = 0.14; among others). This pattern persisted across various iterations, further reinforcing the absence of a causal association.

**Figure 2.**
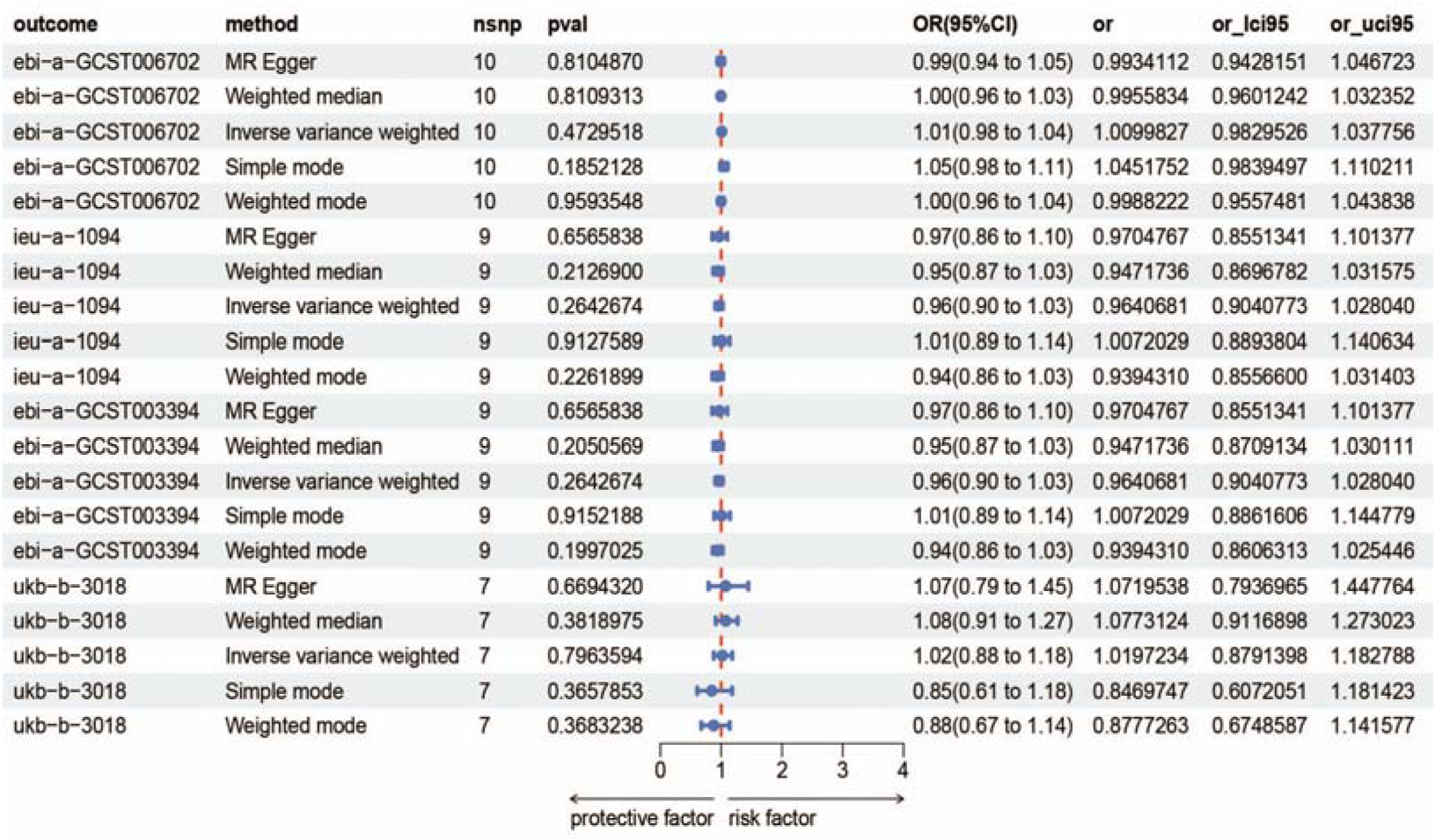
Genetic causal associations between Klotho and human lifespan based on pQTLs-linked MR analysis. The inverse variance weighted method is considered the main method.

In the realm of sensitivity testing, our analysis remained unperturbed by the presence of pleiotropic effects, as evidenced by the results presented in Supplementary Figure 1. Additionally, no single SNP emerged as a sole determinant capable of swaying the overall outcome, underscoring the robustness of our findings (Supplementary Figure 2).

The MR-Egger Intercept analysis, a crucial step in evaluating potential horizontal pleiotropy, yielded no indications of such relationships between α-klotho and human longevity, providing further validation to our conclusions (Supplementary Table 3).

Lastly, the assessment of heterogeneity through the MR-Egger and IVW tests revealed Q-values that did not reach statistical significance (all P values > 0.05), demonstrating the homogeneity and reliability of our analytical approach (Supplementary Table 4). In summary, this comprehensive pQTL-based MR analysis has conclusively demonstrated the absence of a causal relationship between α-klotho and human lifespan.

## Discussion

This pioneering research, for the first time, leverages large-scale genetic datasets to delve into the possible causal link between serum Klotho and human lifespan. Through meticulous analysis, we have arrived at a crucial conclusion: genetically speaking, there is no direct causal connection between Klotho protein and human lifespan. In other words, the levels of Klotho in serum may not serve as a reliable biomarker for predicting human longevity.

Since the discovery of Klotho protein in mice by Kuro-o and colleagues, there has been a growing interest in exploring Klotho’s manifestations in the field of aging. The transmembrane Klotho subtype serves as an essential cofactor in the activation of the FGF signaling pathway, forming a complex with fibroblast growth factor receptor 23(Aczel et al., 2023). Cell surface proteases, known as sheddases, can cleave Klotho, resulting in the production of soluble Klotho(Espuch-Oliver, et al., 2022). This soluble form of Klotho, released into the blood, cerebrospinal fluid, and urine(Jurado-Fasoli, Amaro-Gahete, Gutiérrez, & Castillo, 2019), functions as a hormone and is believed to be associated with longevity in both humans and animals.

Arroyo et al. have highlighted that the expression of Klotho in both mice and humans declines with advancing age, correlating with shorter lifespans(Arroyo, et al., 2022). Conversely, mice with Klotho overexpression exhibit extended lifespans compared to their wild-type counterparts(Kurosu et al., 2005). In humans, higher serum levels of Klotho are likely associated with improved health status and longevity(Espuch-Oliver, et al., 2022). Research indicates serum Klotho levels in humans decrease progressively over time. Additionally, elevated plasma Klotho levels are positively correlated with favorable outcomes, such as a reduced risk of frailty(Shardell et al., 2019). Conversely, lower plasma Klotho levels are associated with negative outcomes, including increased mortality rates in those with deficient Klotho levels(Semba et al., 2011).

Klotho, with its significant influence on human aging and longevity, harbors vast biomarker potential. However, its association with human aging remains controversial. Variations exist, as serum Klotho levels in young males do not consistently exceed those of elders(King, McCormick, Notley, Fujii, & Kenny, 2022). A US cross-sectional study in midlife to older adults uncovered a U-shaped link between serum Klotho and phenotypic age acceleration(Guan, Ma, & Wu, 2023), implying a complex interplay: below a certain level, Klotho promotes aging deceleration, while above, it may accelerate aging. Thus, the Klotho-aging relationship transcends simple direct correlation.

## Conclusions

Collectively, although existing research has proposed Klotho as a potential marker for human lifespan, enabling quantitative predictions(Paquette et al., 2023), clinical randomized trials have yet to confirm this notion. Our innovative Mendelian randomization approach, which leverages natural genetic variation to investigate causality, found no direct genetic causal link between Klotho and human lifespan. This finding underscores the complexity of aging mechanisms and highlights the need for further research to understand the indirect roles Klotho may play in regulating lifespan, potentially through signaling pathways related to aging and disease. Our findings challenge the notion that Klotho levels alone can serve as a definitive predictor of human longevity and encourage further exploration of novel biomarkers and mechanisms that may more accurately gauge human lifespan.

## Data Availability

The datasets analyzed in the current study are available in the open repository as described in the main text. Relevant data or materials can also be obtained from the corresponding author upon reasonable request.

## Acknowledgments

We thank the IEU database for providing a platform for GWAS data querying and downloading.

## Consent for publication

Not applicable

## Authors’ contributions

WY conceived and designed the work. XL developed the experimental methodologies. XL performed the analysis and interpreted the data. WY, XL discussed the results and wrote the paper. And WY supervised the study. ALL authors read and approved the final manuscript.

## Supplementary Figure Legends

**Supplementary Figure 1.**
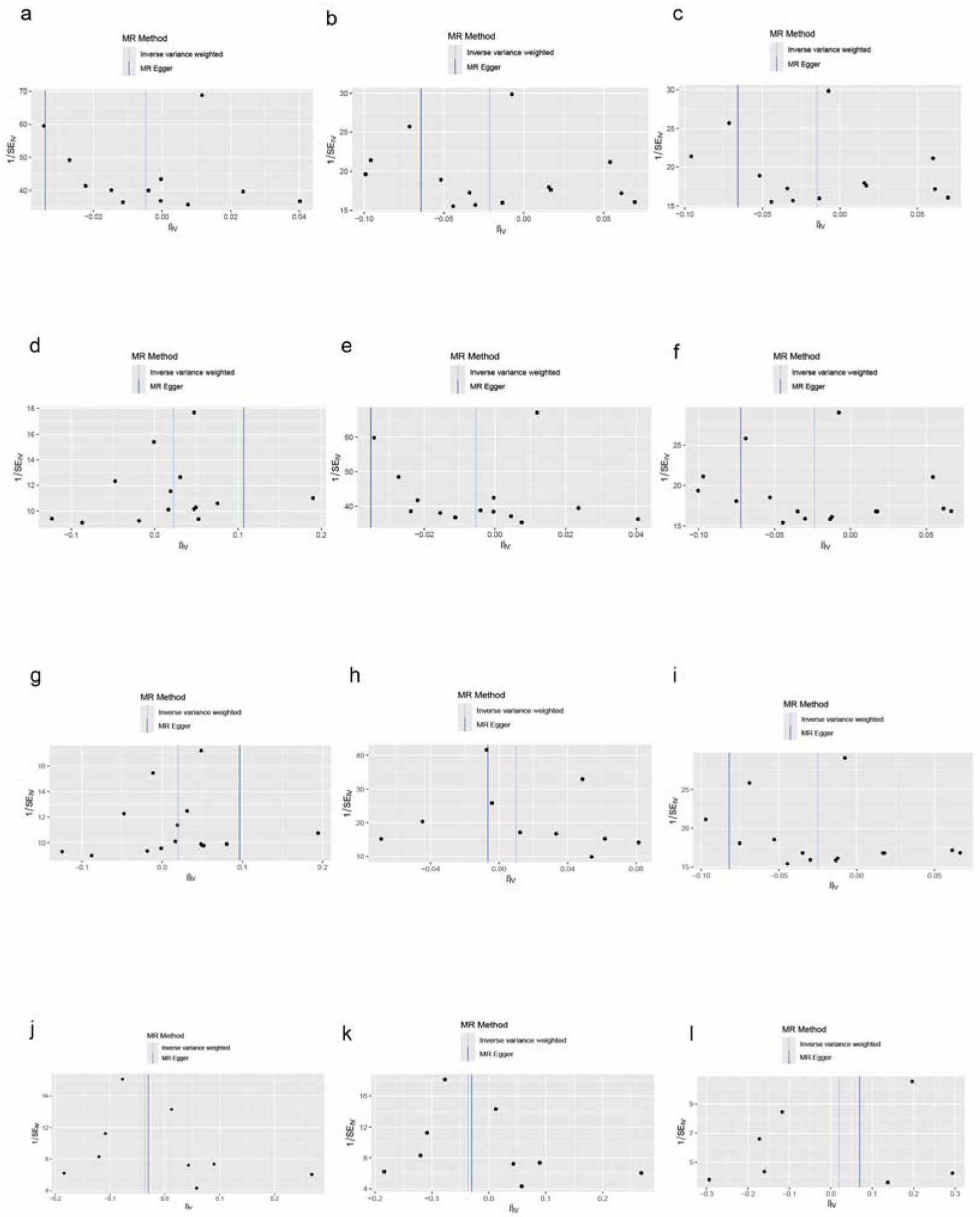
The heterogeneity tests. a-h: The heterogeneity results for GWAS-based MR analysis; i-j: The heterogeneity results for the pQTLs-based MR analysis.

**Supplementary Figure 2.**
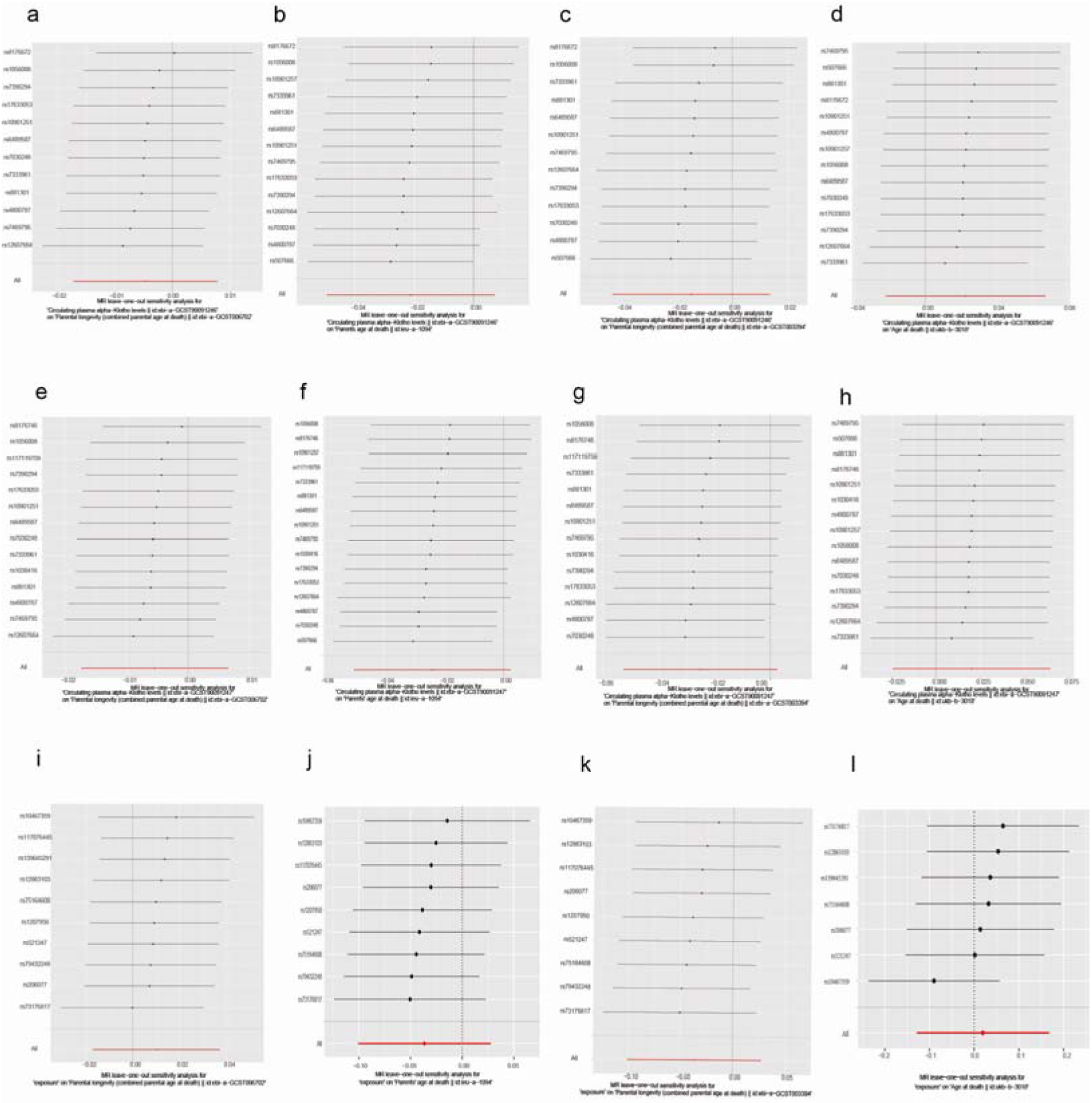
The leave-one-out analysis. a-h: The leave-one-out results for GWAS-based MR analysis; i-j: The leave-one-out results for the pQTLs-based MR analysis.

**Supplementary Table 1.** Detailed information on studies used.

| Dataset | Sexes | Published year | PMID | Sample size | Number of SNPs | Population | Consortium |
| --- | --- | --- | --- | --- | --- | --- | --- |
| **Exposure: Circulating plasma α-Klotho levels** |  |  |  |  |  |  |  |
| ebi-a-GCST90091246<br>( experimental group ) | NA | 2021 | 34542150 | 4 , 376 | 6,418,495 | European | NA |
| ebi-a-GCST90091247<br>( validation group ) | NA | 2021 | 34542150 | 4 , 675 | 6,414,265 | European | NA |
| pQTL(cis) | NA | 2023 | 37794188 | 35 , 559 | 680 | European | NA |
| **Outcome: Age at death** |  |  |  |  |  |  |  |
| ieu-a-1094 | combined | 2016 | 27015805 | 75,244 | 9,583,643 | European | UK Biobank |
| ebi-a-GCST006702 | combined | 2017 | 29227965 | 208, 118 | 11,484,372 | European | NA |
| ebi-a-GCST003394 | combined | 2016 | 27015805 | 45,627 | 9,481,847 | European | NA |
| ukb-b-3018 | combined | 2018 | NA | 11,856 | 9,851,867 | European | MRC-IEU |

**Supplementary Table 2.** Detailed information on genetic instruments associated with exposure.

| Exposure | SNP | Effect_allele | Other_allele | Eaf | Beta | SE | Pval | F-statistic |
| --- | --- | --- | --- | --- | --- | --- | --- | --- |
| Circulating plasma α-Klotho<br>id:ebi-a-GCST90091246 | rs881301 | C | T | 0.4128 | -0.1186 | 0.0212 | 2.23E-08 | 31.29663581 |
|  | rs7469795 | C | T | 0.3014 | 0.1305 | 0.0232 | 1.78E-08 | 31.640625 |
|  | rs10901257 | A | G | 0.2477 | 0.1703 | 0.0247 | 4.96E-12 | 47.53739612 |
|  | rs7390294 | G | A | 0.8385 | -0.1861 | 0.0287 | 9.42E-11 | 42.04641309 |
|  | rs8176672 | T | C | 0.0718 | 0.4061 | 0.0407 | 2.11E-23 | 99.55822854 |
|  | rs507666 | A | G | 0.2144 | -0.2043 | 0.0262 | 5.66E-15 | 60.80428005 |
|  | rs10901251 | C | A | 0.1723 | 0.1628 | 0.0284 | 9.49E-09 | 32.86034517 |
|  | rs7030248 | A | G | 0.349 | 0.1273 | 0.0221 | 7.91E-09 | 33.1796851 |
|  | rs1056008 | C | T | 0.2682 | 0.1835 | 0.0239 | 1.80E-14 | 58.94898549 |
|  | rs6489587 | G | A | 0.7701 | -0.1595 | 0.0253 | 2.93E-10 | 39.74480151 |
|  | rs7333961 | A | G | 0.0462 | -0.3271 | 0.0512 | 1.73E-10 | 40.81512833 |
|  | rs17633053 | T | C | 0.0721 | 0.2712 | 0.0428 | 2.42E-10 | 40.15058084 |
|  | rs4800787 | C | T | 0.7103 | 0.1412 | 0.0233 | 1.42E-09 | 36.72464035 |
|  | rs12607664 | T | G | 0.3161 | 0.2425 | 0.0224 | 2.28E-27 | 117.1999562 |
| Circulating plasma α-Klotho<br>id:ebi-a-GCST90091247 | rs881301 | C | T | 0.4105 | -0.1177 | 0.0205 | 9.76E-09 | 32.96440214 |
|  | rs7030248 | A | G | 0.3472 | 0.1334 | 0.0214 | 4.71E-10 | 38.85832824 |
|  | rs10901251 | C | A | 0.1718 | 0.1651 | 0.0275 | 1.99E-09 | 36.04364959 |
|  | rs8176746 | T | G | 0.0717 | 0.407 | 0.0395 | 6.65E-25 | 106.1682423 |
|  | rs7390294 | G | A | 0.8365 | -0.1742 | 0.0277 | 3.15E-10 | 39.54911442 |
|  | rs507666 | A | G | 0.2179 | -0.2034 | 0.0252 | 6.73E-16 | 65.14795918 |
|  | rs10901257 | A | G | 0.2476 | 0.1679 | 0.0238 | 1.90E-12 | 49.76768943 |
|  | rs7469795 | C | T | 0.3014 | 0.1294 | 0.0224 | 7.19E-09 | 33.37125319 |
|  | rs117119759 | A | G | 0.0228 | 0.4193 | 0.0766 | 4.46E-08 | 29.96347545 |
|  | rs6489587 | G | A | 0.7687 | -0.1554 | 0.0245 | 2.17E-10 | 40.23183673 |
|  | rs1056008 | C | T | 0.2688 | 0.1812 | 0.0232 | 6.32E-15 | 61.00148633 |
|  | rs7333961 | A | G | 0.0465 | -0.3198 | 0.0494 | 9.37E-11 | 41.90858726 |
|  | rs17633053 | T | C | 0.0735 | 0.2584 | 0.0411 | 3.30E-10 | 39.52768454 |
|  | rs1030416 | A | G | 0.3255 | 0.1304 | 0.0232 | 1.88E-08 | 31.5921522 |
|  | rs12607664 | T | G | 0.3172 | 0.2355 | 0.0217 | 1.55E-27 | 117.7775064 |
|  | rs4800787 | C | T | 0.7118 | 0.141 | 0.0225 | 3.97E-10 | 39.27111111 |
| pQTLs of<br>α-Klotho | rs206077 | A | C | 0.19888 | -0.0599 | 0.010638 | 1.79E-08 | 31.70543184 |
|  | rs1207950 | A | G | 0.05983 | -0.0979 | 0.017557 | 2.46E-08 | 31.0931531 |
|  | rs139645291 | T | G | 0.05479 | 0.1218 | 0.018092 | 1.67E-11 | 45.32328904 |
|  | rs79432248 | G | A | 0.02274 | -0.1896 | 0.027564 | 6.05E-12 | 47.31427432 |
|  | rs10467359 | G | A | 0.04031 | -0.3185 | 0.020808 | 6.90E-53 | 234.2923967 |
|  | rs73176817 | C | T | 0.10778 | 0.2011 | 0.013392 | 5.74E-51 | 225.4931014 |
|  | rs521247 | G | A | 0.21378 | -0.0638 | 0.010242 | 4.69E-10 | 38.80358172 |
|  | rs12863103 | T | C | 0.3042 | 0.0932 | 0.009068 | 8.85E-25 | 105.6352348 |
|  | rs75164608 | C | G | 0.05417 | -0.125 | 0.018916 | 3.89E-11 | 43.66781033 |
|  | rs117076445 | A | G | 0.0132 | -0.2048 | 0.037126 | 3.46E-08 | 30.43011432 |

Eaf , effect allele frequency;
Beta , the effect size;
SE , standard error;
Pval , the P-value for the SNP's association.
The genetic instrumental variables were filtered with genome-wide significance through $P < 5e-08$ , and removed Linkage disequilibrium (LD) using a clumping $r^2$ cutoff of 0.3 within a 100Mb window, using the 1000 Genomes Project Phase 3 (EUR) as the reference panel.
the F-statistic for each SNP was calculated as follows: $F = (BATA/SE)^2$ .

**Supplementary Table 3.** The results of pleiotropy test.

| Exposure | Outcome | Egger_intercept | SE | P for pleiotropy |
| --- | --- | --- | --- | --- |
| Circulating plasma α-Klotho levels<br>id:ebi-a-GCST90091246 | ebi-a-GCST006702 | 0.0057902 | 0.00345369<br>6 | 0.124569267 |
|  | ieu-a-1094 | 0.008433005 | 0.00843450<br>5 | 0.337131695 |
|  | ebi-a-GCST003394 | 0.010106476 | 0.00798714<br>2 | 0.231894583 |
|  | ukb-b-3018 | -0.016538333 | 0.01339350<br>3 | 0.240536509 |
| Circulating plasma α-Klotho levels<br>id:ebi-a-GCST90091247 | ebi-a-GCST006702 | 0.005832602 | 0.00311668<br>1 | 0.085858952 |
|  | ieu-a-1094 | 0.00942186 | 0.00713505<br>5 | 0.207848471 |
|  | ebi-a-GCST003394 | 0.01115751 | 0.00718648<br>3 | 0.146489736 |
|  | ukb-b-3018 | -0.014442176 | 0.01298397<br>2 | 0.286152417 |
| pQTLs of α-Klotho | ebi-a-GCST006702 | 0.002532932 | 0.00349053<br>7 | 0.488730849 |
|  | ieu-a-1094 | -0.001022864 | 0.00843679<br>5 | 0.906908959 |
|  | ebi-a-GCST003394 | -0.001022864 | 0.00843679<br>5 | 0.906908959 |
|  | ukb-b-3018 | -0.007475786 | 0.01941886<br>6 | 0.716087341 |
| P values for pleiotropy were derived from MR-Egger test and P < 0.05 indicates a possible pleiotropic effect.<br>SE , standard error. |  |  |  |  |

**Supplementary Table 4.**
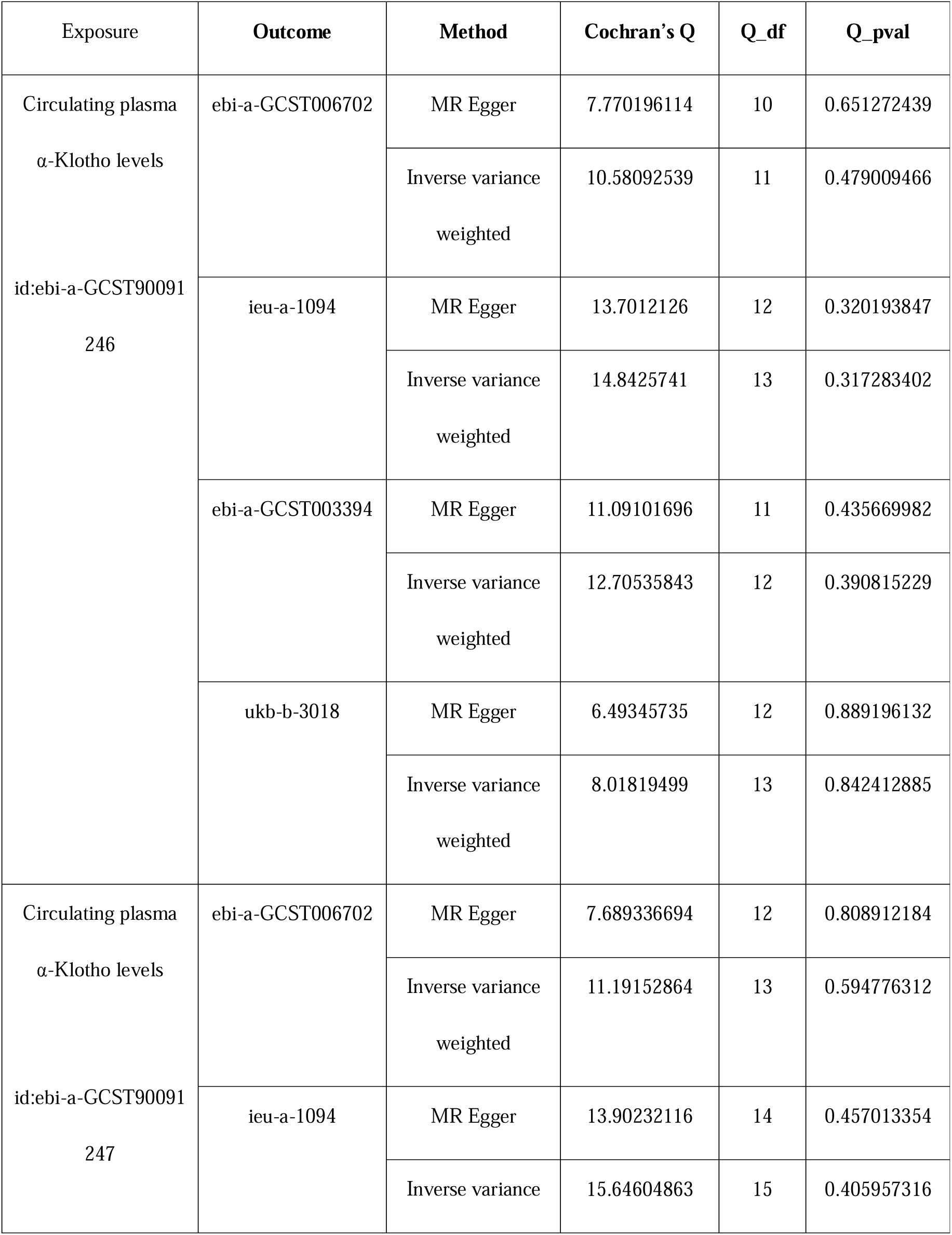
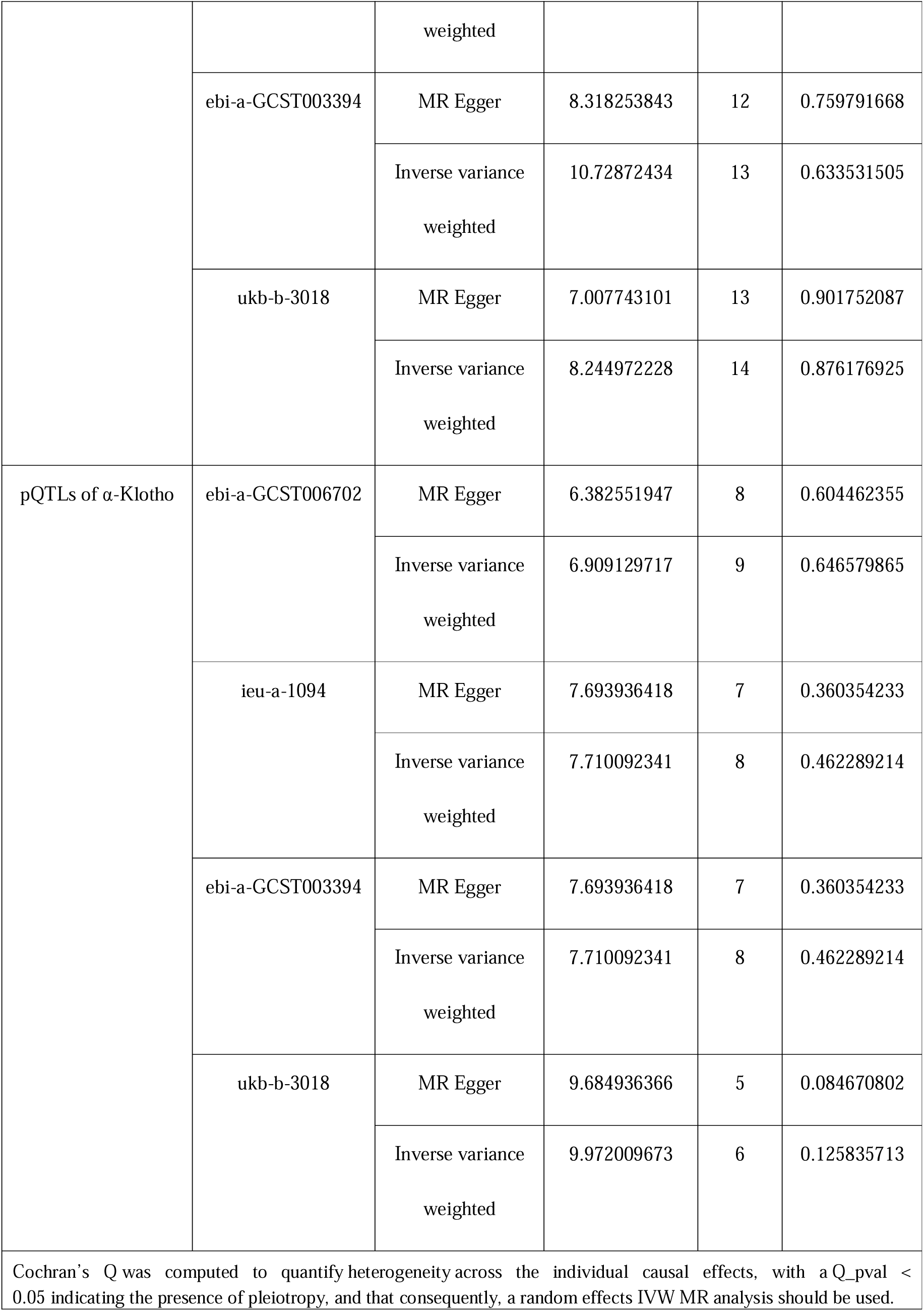
The results of heterogeneity test.

